# Association of Angiotensin-Converting Enzyme Inhibitors and Angiotensin Receptor Blockers with the Risk of Hospitalization and Death in Hypertensive Patients with Coronavirus Disease-19

**DOI:** 10.1101/2020.05.17.20104943

**Authors:** Rohan Khera, Callahan Clark, Yuan Lu, Yinglong Guo, Sheng Ren, Brandon Truax, Erica S Spatz, Karthik Murugiah, Zhenqiu Lin, Saad B Omer, Deneen Vojta, Harlan M Krumholz

## Abstract

**Background:** Whether angiotensin-converting enzyme (ACE) Inhibitors and angiotensin receptor blockers (ARBs) mitigate or exacerbate SARS-CoV-2 infection remains uncertain. In a national study, we evaluated the association of ACE inhibitors and ARB with coronavirus disease-19 (COVID-19) hospitalization and mortality among individuals with hypertension.

**Methods:** Among Medicare Advantage and commercially insured individuals, we identified 2,263 people with hypertension, receiving ≥1 antihypertensive agents, and who had a positive outpatient SARS-CoV-2 test (outpatient cohort). In a propensity score-matched analysis, we determined the association of ACE inhibitors and ARBs with the risk of hospitalization for COVID-19. In a second study of 7,933 individuals with hypertension who were hospitalized with COVID-19 (inpatient cohort), we tested the association of these medications with in-hospital mortality. We stratified all our assessments by insurance groups.

**Results:** Among individuals in the outpatient and inpatient cohorts, 31.9% and 29.8%, respectively, used ACE inhibitors and 32.3% and 28.1% used ARBs. In the outpatient study, over a median 30.0 (19.0 - 40.0) days after testing positive, 12.7% were hospitalized for COVID-19. In propensity score-matched analyses, neither ACE inhibitors (HR, 0.77 [0.53, 1.13], P = 0.18), nor ARBs (HR, 0.88 [0.61, 1.26], P = 0.48), were significantly associated with risk of hospitalization. In analyses stratified by insurance group, ACE inhibitors, but not ARBs, were associated with a significant lower risk of hospitalization in the Medicare group (HR, 0.61 [0.41, 0.93], P = 0.02), but not the commercially insured group (HR: 2.14 [0.82, 5.60], P = 0.12; P-interaction 0.09). In the inpatient study, 14.2% died, 59.5% survived to discharge, and 26.3% had an ongoing hospitalization. In propensity score-matched analyses, neither use of ACE inhibitor (0.97 [0.81, 1.16]; P = 0.74) nor ARB (1.15 [0.95, 1.38]; P = 0.15) was associated with risk of in-hospital mortality, in total or in the stratified analyses.

**Conclusions:** The use of ACE inhibitors and ARBs was not associated with the risk of hospitalization or mortality among those infected with SARS-CoV-2. However, there was a nearly 40% lower risk of hospitalization with the use of ACE inhibitors in the Medicare population. This finding merits a clinical trial to evaluate the potential role of ACE inhibitors in reducing the risk of hospitalization among older individuals, who are at an elevated risk of adverse outcomes with the infection.

## BACKGROUND

Whether the use of angiotensin-converting enzyme (ACE) inhibitors and angiotensin receptor blockers (ARBs) mitigates or exacerbates SARS-CoV-2 infection remains unknown.^1^ Experts have postulated, based on the effect of the drugs and the mechanism of virus entry, that ACE inhibitors and ARBs could be beneficial, harmful or have no effect on people infected with SARS-CoV-2.^1–3^ Evaluations of the mechanism of action of these drugs also suggests differences between the outcomes of patients with ACE inhibitors and ARBs.^4^ There is evidence from randomized controlled trials predating coronavirus disease-19 (COVID-19) suggesting a decrease in risk of all-cause pneumonia with ACE inhibitors, an effect not observed with ARBs.^5^

Recent studies that have focused on the association of ACE inhibitors and ARBs with the risk of mortality among patients hospitalized with COVID-19 suggest that these drugs are not harmful,^6^ with some suggesting that ACE inhibitors may reduce this risk of in-hospital death.^1,7–9^ These studies were limited by their designs, which lacked an active comparator.^4,7^ Moreover, no large national study has addressed the association of these drugs with outcomes among individuals in the outpatient setting infected with SARS-CoV-2. The issue is important because these drugs are widely available and inexpensive and, if beneficial, could modify disease course and improve outcomes. Alternatively, if they increase risk, they could be compounding the harm caused by the virus.

Accordingly, we sought to conduct a large, national study of the association of ACE inhibitors and ARBs with outcomes in patients with hypertension. We specifically evaluated the association of the use of ACE inhibitors and ARBs among patients with hypertension so that we could have an active comparator, other antihypertensive agents. Also, to provide information about the association in inpatients, we conducted a study of the association of ACE inhibitors and ARBs on mortality among people with hypertension who were hospitalized with COVID-19. We stratified all our assessments by insurance groups due to substantial differences between the two populations.

## METHODS

### Overview

We conducted 2 studies of patients with hypertension – the first study included individuals who tested positive for SARS-CoV-2 as an outpatient and the second included patients hospitalized with COVID-19. In addition to a diagnosis of hypertension, we prespecified our study population to include individuals that were receiving at least 1 antihypertensive agent. Further, to account for medical comorbidities, we created robust propensity score matched cohorts of patients treated with ACE inhibitors, ARBs and other antihypertensive agents. We evaluated the success of our matching algorithms through explicit assessments of covariate balance across all comparisons and evaluation of exposure groups on falsification endpoints.

Due to systematic differences among enrollees in Medicare Advantage and commercial insurance programs, and the enhanced risk of adverse outcomes with COVID-19 among older individuals who are overrepresented in Medicare,^10–13^ we evaluated the association of these drugs with outcomes in analyses stratified by insurance groups.

### Data Sources

We used de-identified administrative claims for Medicare Advantage and commercially insured members in a research database from a single large US health insurance provider. The database contains medical (emergency, inpatient, outpatient) and pharmacy claims for services submitted for third party reimbursement, available as International Classification of Diseases, Tenth Revision, Clinical Modification (ICD-10-CM) and National Drug Codes (NDC) claims, respectively. These claims are aggregated after completion of care encounters and submission of claims for reimbursement.

There were two additional data sources that included information on COVID-19 and could be linked to the claims. First, the limited outpatient testing dataset included information on SARS-CoV-2 test results for members who underwent outpatient testing for SARS-CoV2 at 49 hospital-based, free-standing outpatient and third-party labs in all states across the United States, with over 90% of tests submitted by third-party lab vendors. Second, the inpatient COVID-19 dataset included a daily-updated record of COVID-19 inpatient admissions for all insurance enrollees with claims information, representing those admitted to a hospital with a primary or secondary diagnosis of COVID-19 (**eTable 1**), along with their current disposition (admitted, discharged, transferred, expired, or unknown).

### Study Population

We constructed cohorts of patients for each of the two studies. First, for the outpatient study, we identified outpatients who tested positive for SARS-CoV-2. We included individuals at least 18 years of age, with at least 6 months of enrollment in Medicare Advantage or commercial insurance from January through December 2019 and available claims data, a diagnosis of hypertension in one or more claims and receiving one or more anti-hypertensive agents, and tested positive for SARS-CoV-2 positive in an outpatient setting. between March 6, 2020 and May 3, 2020 (**eFigure 1**). The Medicare Advantage and commercially insured individuals in the study represented all individuals with available claims in the UnitedHealth Groups Clinical Research Database that satisfied the inclusion criteria. We evaluated the association of ACE inhibitors and ARBs, respectively, on the risk of hospitalization in this outpatient cohort compared with patients on other antihypertensive medications.

Second, for the second inpatient study, we identified an inpatient cohort of adults hospitalized with COVID-19. This included all patients (age ≥18 years) with at least 6 months of health insurance enrollment in 2019 with available claims data, a diagnosis of hypertension in one or more claims, were receiving one or more anti-hypertensive agents, and were hospitalized with a principal or secondary diagnosis of COVID-19 between January 5, 2020 and May 10, 2020 (**eFigure 2**). We evaluated the association of ACE inhibitors and ARBs, respectively, on the mortality of hospitalized individuals in this cohort compared with patients on other antihypertensive medications.

For both studies, a diagnosis of hypertension was based on ICD-10 codes (**eTable 1**), and drug treatment for hypertension was defined by the receipt of one or more agents included in the 2017 American Heart Association hypertension guidelines.^14^ These include first-line agents of ACE inhibitors, ARBs, thiazide and thiazide-like diuretics, and dihydropyridine and non-dihydropyridine calcium channel blockers, as well as second-line agents of beta-adrenergic antagonists, alpha blockers, centrally acting alpha agonists, loop diuretics, potassium sparing diuretics, mineralocorticoid receptor antagonists, and direct vasodilators (individual drugs listed in **eTable 2**). The information on the latter was derived from pharmacy claims and was defined by a cumulative supply greater than 30 days between July 1, 2019 and December 31, 2019.

### Study Exposures

We identified 2 mutually exclusive exposure groups (1) Patients receiving ACE inhibitors, and (2) those receiving ARB, with or without other agents. We used an active comparator for these analyses that included all remaining patients with a diagnosis of hypertension who received one or more anti-hypertensive agents from drug classes other than ACE inhibitor or ARB. These agents include all first and second line anti-hypertensive agents based on the 2017 AHA guidelines for hypertension,^14^ with the use of individual agents defined by one or more pharmacy claims for at least 30 days of medication supply for the agent between July and December 2019. In sensitivity analyses, we restricted the control group to individuals receiving at least one first line anti-hypertensive agent from drug classes other than ACE inhibitor or ARB.

### Study Covariates

We combined information from inpatient and outpatient claims in 2019 to identify potential confounders of the association of the ACE inhibitors and ARB use and patient outcomes. We included patient age, sex, race, conditions that would represent potential indications for selective use of ACE inhibitors or ARBs (diabetes, myocardial infarction, heart failure and chronic kidney disease), and each of the additional comorbidities included in the Charlson Comorbidity Index (peripheral vascular disease, cerebrovascular disease, hemi- or paraplegia, dementia, chronic pulmonary disease, rheumatologic disease, diabetes with chronic complications, malignancy, metastatic solid tumor, mild liver disease, moderate-to-severe liver disease, acquired immunodeficiency syndrome or human immunodeficiency virus). The ICD-10 codes corresponding to these variables are included in **eTable 1** (appendix). Race was available only for Medicare Advantage members. We included information on the total number of anti-hypertensive agents prescribed to patients using pharmacy claims. Finally, regional clustering of cases was identified using location of sites submitting claims for laboratory testing for SARS-CoV-2 and hospital site for in-hospital outcomes.

### Study Outcomes

In the outpatient study, the primary outcome was inpatient hospitalization for COVID-19, defined as a hospitalization with a principal or a secondary diagnosis of COVID-19 in a linked inpatient dataset (**eTable 1**). We assessed mortality during this inpatient hospitalization as a secondary outcome.

In the inpatient study, the primary outcome was in-hospital mortality. In addition, we evaluated a secondary composite outcome of death or discharge to hospice and hospital length of stay.

### Propensity score matching

In both outpatient and inpatient studies, we created propensity score-matched cohorts of patients with hypertension, treated with ACE inhibitors, ARBs or other antihypertensive medications. For this, we constructed a non-parsimonious multivariable logistic regression model with receipt of ACE inhibitors, ARB or other antihypertensive as the dependent variable. For example, we modeled the receipt ACE inhibitor or another other anti-hypertensive (excluding ARB) to determine each patient’s likelihood of receiving these agents based on their measured clinical characteristics. We applied this strategy to different pairs of treatment comparisons (ACE inhibitor vs others, ARB vs others, and ACE inhibitor vs ARB). Briefly, we performed 1:1 matching between recipients based on propensity scores with a caliper width of one-tenth of the standard deviation of the logit of the propensity score.^4^ We pursued 100 iterations to find the lowest mean absolute standardized difference among matched variables. We matched our cohorts on age, gender, race, insurance type, conditions that may lead to selective use of ACE inhibitors and ARBs (i.e., diabetes, myocardial infarction, heart failure and chronic kidney disease), each of the comorbidities in the Charlson Comorbidity Index, and the number of anti-hypertensive agents used for the patient. To account for regional clustering of care practices and response to the COVID-19 pandemic, we explicitly accounted for census region of lab testing site or inpatient facility in our models.

We evaluated the performance of propensity score matching using several strategies. First, we assessed the propensity score distributions in the unmatched and matched cohorts and calculated an equipoise metric to summarize the degree of overlap in characteristics of patients receiving these drugs.^15,16^ This represents the proportion of individuals in the unmatched groups that had a propensity score between 0.3 and 0.7, representing a state of equipoise between the two drugs. A value greater than 0.5 implies two drugs are in empirical equipoise, with a higher a value indicating a lower likelihood of confounding by indication.^15^ Next, we evaluated the standardized difference between matched covariates before and after propensity score matching. Specifically, we evaluated whether our matching algorithm achieved a standardized difference of <10% between matched cohort suggestive of adequately matched groups.^16,17^ Second, we evaluated the success of our matching algorithm using a priori defined negative control outcomes. We chose two negative control or falsification endpoints – claims for gastroesophageal reflux disease and ingrown nail - that are unlikely to be affected by the treatment assignment and a directional effect would represent covariate imbalance.^16^ Third, we evaluated whether our propensity matched cohort were also adequately matched on other therapeutic classes for non-antihypertensive agents that were not directly included in the matching algorithm. These strategies were designed to evaluate the potential for residual confounding after creating propensity score matched cohorts. Finally, we evaluated our observations for robustness by assessing treatment effects in 100 iterations of the propensity score matching algorithm, evaluating whether our findings were consistent across these iterations that varied on the degree of matching of individual covariates.

### Statistical analyses

We describe differences between patients treated with ACE inhibitors and ARBs compared with other anti-hypertensive agents, and between those treated with ACE inhibitors using Chi square test for categorical variables and t-test for continuous variables. As the duration of follow up was expected to vary across individuals in both outpatient and inpatient COVID-19 cohorts, we evaluated their effects in time-to-event analyses with Cox-Proportional Hazards models in both unadjusted and propensity score-matched cohorts. To reduce bias from residual differences in matched covariates in our evaluation of patient outcomes, we included the covariates included in our propensity score matching algorithm as independent variables in these models.^18^ We repeated these analyses without this additional covariate adjustment.

For the outpatient study, the index date was represented by the day of positive COVID-19 test as an outpatient, the period of the study was measured in days from the positive COVID-19, and the outcome of interest was hospitalization. For the inpatient study, the index date was represented by the first day of hospitalization with COVID-19, the period of the study was measured in days from admission, and the outcome of interest was death. Since hospitalization could end with either patient’s death or being discharged alive, we created a cause-specific Cox Proportional Hazards model, which is a competing risk analysis.^19–21^ Both analyses were censored at the end of the observation period on May 10, 2020.

Given systematic differences between patients enrolled in Medicare Advantage and commercial insurance, particularly with older age, higher comorbidity burden, and higher risk of COVID-19 complications among Medicare advantage enrollees, we evaluated quantitative and qualitative interactions between insurance type and treatment groups for the assessment of our outcomes. We also created propensity score-matched cohorts within the each of the two insurance subgroups.

Analyses were performed using open source R 3.4.0 (CRAN) and Python 3.8.2. All hypothesis tests were 2-sided, with a level of significance set at 0.05, except for interaction tests where the level of significance was set at 0.10. Given the exploratory nature of study, statistical tests were not adjusted for multiple testing. The Yale Institutional Review Board and the UnitedHealth Group Office of Human Research Affairs exempted this study from other review as all activities were limited to retrospective analysis of de-identified data and accessed in accordance with Health Insurance Portability and Accountability Act regulations.

## RESULTS

### Characteristics of the Outpatient Cohort

Among 6,885 patients who tested positive for SARS-CoV-2 and had at least 6 months of enrollment in Medicare Advantage or commercial insurance, and in pharmacy benefits with their insurance, 2,263 had a diagnosis of hypertension with the use of at least one anti-hypertensive drug (**eFigure 1**). The outpatient study cohort included individuals from 44 states in the US (**eFigure 3**). A total of 1,467 (64.8%) were Medicare Advantage enrollees and 796 (35.2%) of the cohort were commercially insured. The median age of these individuals was 69.0 years and 52.5% were women. Of Medicare Advantage patients, 29.6% were African American. Patients receiving ACE inhibitors or ARBs were more likely to be men, have diabetes with or without chronic complications, and be receiving more than 1 antihypertensive drugs (**Table 1)**. They were less likely to have chronic heart failure or moderate-to-severe renal disease, compared with patients receiving other anti-hypertensive treatments. Patients receiving ACE inhibitors were similar to those receiving ARBs but had a higher rate of dementia and peripheral vascular disease (**Table 1**). We matched 441 patients receiving ACE inhibitors to 441 patients receiving other antihypertensive agents (**eFigure 5**), achieving <10% standardized differences for all covariates (**Figure 1**). Similarly, we matched 412 patients receiving ARB to 412 patients receiving other antihypertensive agents (**eFigure 5**). The equipoise for comparisons of ACE inhibitors to other drugs, and for ACE inhibitors to ARB were above 0.5, but were lower for the ARB comparisons (**Table 3)**.

**Table 1:** Characteristics of Outpatient Study Cohort. The cohort includes patients who had a positive test for SARS-CoV-2 in the outpatient setting.

| Variable | Overall | Antihypertensive Drug Cohorts |  |  | p-value |  |  |
| --- | --- | --- | --- | --- | --- | --- | --- |
|  |  | ACE inhibitor | ARB | Other | ACE inhibitor vs Other | ARB vs Other | ACE inhibitor vs ARB |
| Number of Patients | 2263 (100.0%) | 722 (100.0%) | 731 (100.0%) | 810 (100.0%) |  |  |  |
| Age, median (IQR) | 69.0 (59.0 - 78.0) | 68.0 (57.0 - 76.0) | 69.0 (59.0 - 76.0) | 71.0 (60.2 - 80.0) | <.001 | <.001 | 0.09 |
| **Age range** |  |  |  |  |  |  |  |
| 18 to 30 y | 10 (0.4%) | 4 (0.6%) | 3 (0.4%) | 3 (0.4%) | 0.88 | 0.78 | 0.99 |
| 31 to 40 y | 63 (2.8%) | 25 (3.5%) | 14 (1.9%) | 24 (3.0%) | 0.68 | 0.25 | 0.10 |
| 41 to 50 y | 171 (7.6%) | 69 (9.6%) | 56 (7.7%) | 46 (5.7%) | 0.01 | 0.14 | 0.23 |
| 51 to 60 y | 408 (18.0%) | 138 (19.1%) | 140 (19.2%) | 130 (16.0%) | 0.13 | 0.13 | 0.96 |
| 61 to 70 y | 570 (25.2%) | 189 (26.2%) | 203 (27.8%) | 178 (22.0%) | 0.06 | 0.01 | 0.53 |
| 71 to 80 y | 606 (26.8%) | 185 (25.6%) | 194 (26.5%) | 227 (28.0%) | 0.32 | 0.55 | 0.74 |
| > 80 y | 435 (19.2%) | 112 (15.5%) | 121 (16.6%) | 202 (24.9%) | <.001 | <.001 | 0.64 |
| Female sex | 1189 (52.5%) | 318 (44.0%) | 392 (53.6%) | 479 (59.1%) | <.001 | 0.03 | <.001 |
| Medicare Advantage | 1467 (64.8%) | 434 (60.1%) | 452 (61.8%) | 581 (71.7%) | <.001 | <.001 | 0.54 |
| **Location** |  |  |  |  |  |  |  |
| Urban | 1059 (46.8%) | 333 (46.1%) | 336 (46.0%) | 390 (48.1%) | 0.46 | 0.42 | 0.99 |
| Rural | 434 (19.2%) | 130 (18.0%) | 142 (19.4%) | 162 (20.0%) | 0.35 | 0.83 | 0.53 |
| Suburban | 747 (33.0%) | 244 (33.8%) | 248 (33.9%) | 255 (31.5%) | 0.36 | 0.33 | 1.00 |
| Unknown | 23 (1.0%) | 15 (2.1%) | 5 (0.7%) | 3 (0.4%) | <.001 | 0.62 | 0.04 |
| **Race \*** |  |  |  |  |  |  |  |
| Caucasian | 863 (38.1%) | 270 (37.4%) | 250 (34.2%) | 343 (42.3%) | 0.05 | <.001 | 0.22 |
| African American | 434 (19.2%) | 113 (15.7%) | 131 (17.9%) | 190 (23.5%) | <.001 | 0.01 | 0.28 |
| Hispanic | 69 (3.0%) | 21 (2.9%) | 25 (3.4%) | 23 (2.8%) | 0.94 | 0.61 | 0.68 |
| Asian | 46 (2.0%) | 15 (2.1%) | 19 (2.6%) | 12 (1.5%) | 0.49 | 0.17 | 0.63 |
| Native American | 2 (0.1%) | 0 (0.0%) | 0 (0.0%) | 2 (0.2%) | 0.53 | 0.52 |  |
| Other | 36 (1.6%) | 10 (1.4%) | 18 (2.5%) | 8 (1.0%) | 0.63 | 0.04 | 0.19 |
| Unknown | 813 (35.9%) | 293 (40.6%) | 288 (39.4%) | 232 (28.6%) | <.001 | <.001 | 0.68 |
| **Geography** |  |  |  |  |  |  |  |
| **Region of Test Site** |  |  |  |  |  |  |  |
| Northeast | 847 (37.4%) | 241 (33.4%) | 242 (33.1%) | 364 (44.9%) | <.001 | <.001 | 0.96 |
| South | 711 (31.4%) | 229 (31.7%) | 275 (37.6%) | 207 (25.6%) | 0.01 | <.001 | 0.02 |
| Midwest | 175 (7.7%) | 64 (8.9%) | 53 (7.3%) | 58 (7.2%) | 0.26 | 0.98 | 0.30 |
| West | 202 (8.9%) | 81 (11.2%) | 64 (8.8%) | 57 (7.0%) | 0.01 | 0.25 | 0.14 |
| Unknown | 328 (14.5%) | 107 (14.8%) | 97 (13.3%) | 124 (15.3%) | 0.85 | 0.29 | 0.44 |
| **State of Test Site** |  |  |  |  |  |  |  |
| New York | 230 (10.2%) | 54 (7.5%) | 79 (10.8%) | 97 (12.0%) | <.001 | 0.52 | 0.03 |
| New Jersey | 303 (13.4%) | 86 (11.9%) | 93 (12.7%) | 124 (15.3%) | 0.06 | 0.17 | 0.70 |
| Connecticut | 136 (6.0%) | 43 (6.0%) | 36 (4.9%) | 57 (7.0%) | 0.45 | 0.10 | 0.45 |
| Georgia | 183 (8.1%) | 58 (8.0%) | 67 (9.2%) | 58 (7.2%) | 0.58 | 0.18 | 0.50 |
| Florida | 124 (5.5%) | 38 (5.3%) | 58 (7.9%) | 28 (3.5%) | 0.11 | 0.00 | 0.05 |
| Other | 959 (42.4%) | 336 (46.5%) | 301 (41.2%) | 322 (39.8%) | 0.01 | 0.61 | 0.04 |
| Unknown | 328 (14.5%) | 107 (14.8%) | 97 (13.3%) | 124 (15.3%) | 0.85 | 0.29 | 0.44 |
| **Comorbid Conditions** |  |  |  |  |  |  |  |
| Diabetes without complications | 911 (40.3%) | 320 (44.3%) | 321 (43.9%) | 270 (33.3%) | <.001 | <.001 | 0.92 |
| Myocardial infarction | 81 (3.6%) | 16 (2.2%) | 20 (2.7%) | 45 (5.6%) | <.001 | 0.01 | 0.64 |
| Chronic heart failure | 326 (14.4%) | 72 (10.0%) | 99 (13.5%) | 155 (19.1%) | <.001 | <.001 | 0.04 |
| Chronic pulmonary disease | 410 (18.1%) | 100 (13.9%) | 139 (19.0%) | 171 (21.1%) | <.001 | 0.34 | 0.01 |
| Peptic ulcer disease | 19 (0.8%) | 6 (0.8%) | 8 (1.1%) | 5 (0.6%) | 0.85 | 0.46 | 0.81 |
| AIDS | 22 (1.0%) | 5 (0.7%) | 11 (1.5%) | 6 (0.7%) | 0.85 | 0.23 | 0.22 |
| Rheumatologic disease | 120 (5.3%) | 28 (3.9%) | 40 (5.5%) | 52 (6.4%) | 0.03 | 0.50 | 0.19 |
| Diabetes, chronic complications | 625 (27.6%) | 225 (31.2%) | 210 (28.7%) | 190 (23.5%) | <.001 | 0.02 | 0.34 |
| Metastatic cancer | 20 (0.9%) | 5 (0.7%) | 5 (0.7%) | 10 (1.2%) | 0.41 | 0.40 | 0.77 |
| Hemiplegia or paraplegia | 92 (4.1%) | 29 (4.0%) | 15 (2.1%) | 48 (5.9%) | 0.11 | 0.00 | 0.04 |
| Liver disease, mild | 106 (4.7%) | 28 (3.9%) | 34 (4.7%) | 44 (5.4%) | 0.19 | 0.56 | 0.55 |
| Solid tumor without metastases | 181 (8.0%) | 41 (5.7%) | 61 (8.3%) | 79 (9.8%) | <.001 | 0.38 | 0.06 |
| Liver disease, moderate to severe | 5 (0.2%) | 2 (0.3%) | 1 (0.1%) | 2 (0.2%) | 0.70 | 0.93 | 0.99 |
| Dementia | 250 (11.0%) | 60 (8.3%) | 43 (5.9%) | 147 (18.1%) | <.001 | <.001 | 0.09 |
| Peripheral vascular disease | 467 (20.6%) | 122 (16.9%) | 121 (16.6%) | 224 (27.7%) | <.001 | <.001 | 0.92 |
| Renal failure, moderate to severe | 359 (15.9%) | 100 (13.9%) | 93 (12.7%) | 166 (20.5%) | <.001 | <.001 | 0.58 |
| Cerebrovascular disease | 289 (12.8%) | 73 (10.1%) | 83 (11.4%) | 133 (16.4%) | <.001 | 0.01 | 0.50 |
| Charlson Score, median (IQR) | 2.0 (0.0 - 3.0) | 1.0 (0.0 - 3.0) | 1.0 (0.0 - 3.0) | 2.0 (0.0 - 4.0) | <.001 | <.001 | 0.29 |
| **Drug Therapy** |  |  |  |  |  |  |  |
| **Antihypertensives** |  |  |  |  |  |  |  |
| Beta blockers | 911 (40.3%) | 243 (33.7%) | 265 (36.3%) | 403 (49.8%) | <.001 | <.001 | 0.33 |
| Non-dihydropyridine calcium channel blockers | 99 (4.4%) | 19 (2.6%) | 23 (3.1%) | 57 (7.0%) | <.001 | <.001 | 0.67 |
| Dihydropyridine calcium channel blockers | 813 (35.9%) | 215 (29.8%) | 253 (34.6%) | 345 (42.6%) | <.001 | <.001 | 0.06 |
| Thiazide or thiazide-like diuretics | 709 (31.3%) | 236 (32.7%) | 300 (41.0%) | 173 (21.4%) | <.001 | <.001 | 0.00 |
| Loop diuretics | 328 (14.5%) | 73 (10.1%) | 84 (11.5%) | 171 (21.1%) | <.001 | <.001 | 0.45 |
| Centrally acting alpha agonists | 54 (2.4%) | 8 (1.1%) | 19 (2.6%) | 27 (3.3%) | 0.01 | 0.49 | 0.06 |
| Potassium sparing diuretics | 56 (2.5%) | 15 (2.1%) | 10 (1.4%) | 31 (3.8%) | 0.06 | <.001 | 0.40 |
| Mineralocorticoid Aldosterone Antagonists | 85 (3.8%) | 15 (2.1%) | 28 (3.8%) | 42 (5.2%) | <.001 | 0.25 | 0.07 |
| Renin inhibitors | 0 (0.0%) | 0 (0.0%) | 0 (0.0%) | 0 (0.0%) |  |  |  |
| Alpha adrenergic blocking agents | 40 (1.8%) | 8 (1.1%) | 15 (2.1%) | 17 (2.1%) | 0.18 | 0.91 | 0.22 |
| Direct vasodilators | 99 (4.4%) | 27 (3.7%) | 27 (3.7%) | 45 (5.6%) | 0.12 | 0.11 | 0.93 |
| **Place in Therapy** |  |  |  |  |  |  |  |
| First line | 1964 (86.8%) | 722 (100.0%) | 731 (100.0%) | 511 (63.1%) | <.001 | <.001 |  |
| Second line | 1135 (50.2%) | 290 (40.2%) | 308 (42.1%) | 537 (66.3%) | <.001 | <.001 | 0.48 |
| **Number of antihypertensive classes** |  |  |  |  |  |  |  |
| 1 | 822 (36.3%) | 206 (28.5%) | 148 (20.2%) | 468 (57.8%) | <.001 | <.001 | <.001 |
| 2 | 780 (34.5%) | 271 (37.5%) | 288 (39.4%) | 221 (27.3%) | <.001 | <.001 | 0.50 |
| 3+ | 661 (29.2%) | 245 (33.9%) | 295 (40.4%) | 121 (14.9%) | <.001 | <.001 | 0.01 |
| Number, median (IQR) | 2.0 (1.0 - 3.0) | 2.0 (1.0 - 3.0) | 2.0 (2.0 - 3.0) | 1.0 (1.0 - 2.0) | <.001 | <.001 | <.001 |
| **Other Drug Therapies** |  |  |  |  |  |  |  |
| Statins | 1210 (53.5%) | 418 (57.9%) | 401 (54.9%) | 391 (48.3%) | <.001 | 0.01 | 0.26 |
| Other lipid lowering agents | 113 (5.0%) | 37 (5.1%) | 42 (5.7%) | 34 (4.2%) | 0.46 | 0.20 | 0.68 |
| Oral anticoagulants | 201 (8.9%) | 48 (6.6%) | 58 (7.9%) | 95 (11.7%) | <.001 | 0.02 | 0.40 |
| Insulins | 215 (9.5%) | 77 (10.7%) | 70 (9.6%) | 68 (8.4%) | 0.15 | 0.47 | 0.55 |
| Oral antihyperglycemic agents | 581 (25.7%) | 234 (32.4%) | 217 (29.7%) | 130 (16.0%) | <.001 | <.001 | 0.29 |
| **Follow up** |  |  |  |  |  |  |  |
| Follow up days, median (IQR) | 30.0 (19.0 - 40.0) | 30.0 (19.0 - 40.0) | 31.0 (21.0 - 40.0) | 29.0 (19.0 - 38.0) | 0.35 | 0.01 | 0.13 |
| Test to hospitalization, median (IQR) | 4.0 (2.0 - 7.0) | 4.0 (2.0 - 6.0) | 4.0 (2.0 - 7.0) | 5.0 (3.0 - 7.0) | 0.40 | 0.51 | 0.83 |
| Total hospitalized | 287 (12.7%) | 77 (10.7%) | 93 (12.7%) | 117 (14.4%) | 0.03 | 0.36 | 0.25 |

\*Race unknown in all commercially insured patients

**Figure 1:**
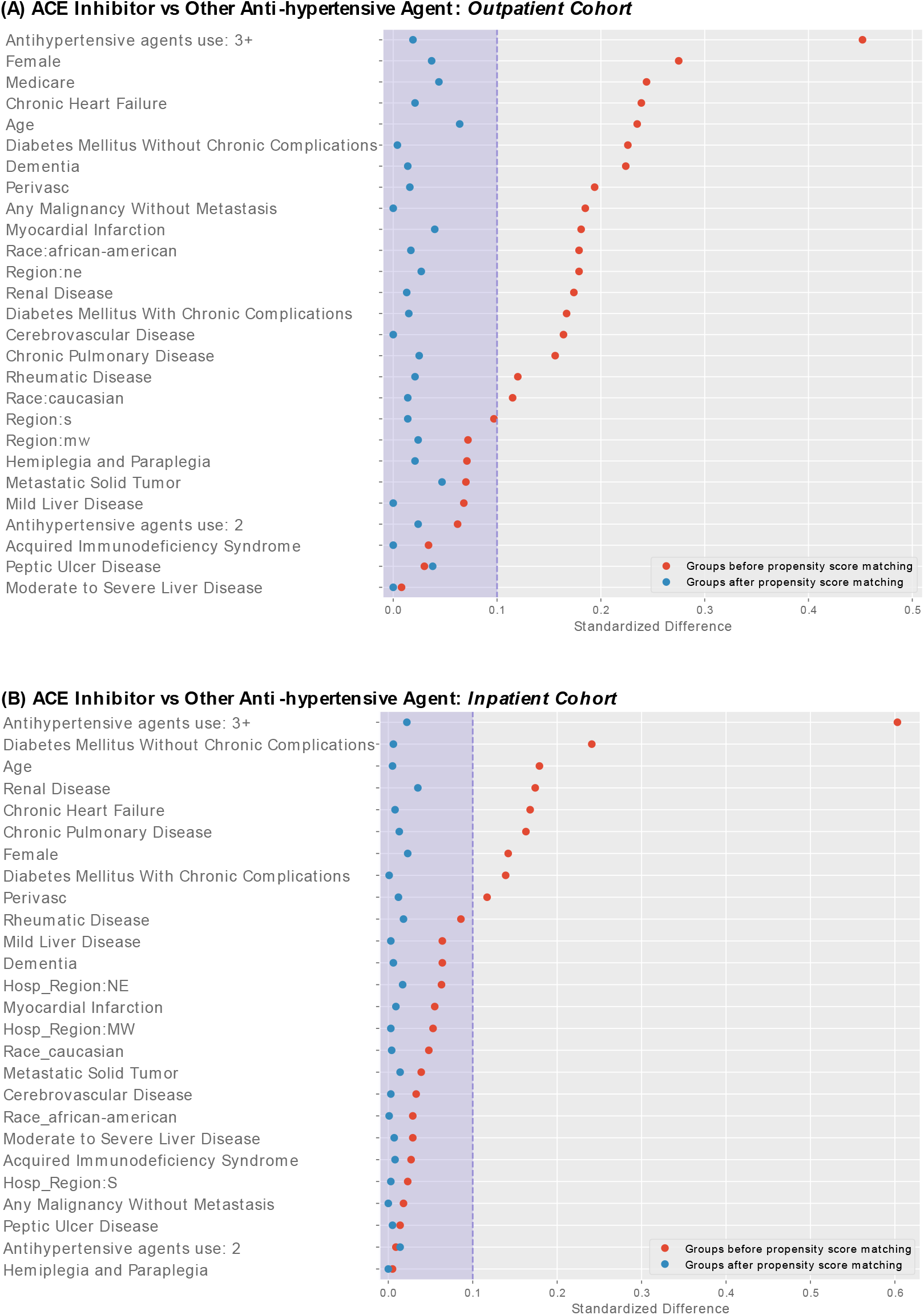
Standardized Differences Between Variables Before and After Propensity Matching.

### Characteristics of the Inpatient Cohort

Among 12,566 patients who were hospitalized for COVID-19 with linked claims data, 7,933 had had a diagnosis of hypertension and had an outpatient prescription for at least one antihypertensive drug (**eFigure 2**). The inpatient cohort included patients from 47 states in the US (**eFigure 3**). Of the included patients, 92.0% were Medicare Advantage enrollees. The median age of hospitalized individuals was 77.0 years, 54.6% were women; 29.9% of Medicare Advantage enrollees were African American. Differences between hospitalized patients treated with ACE inhibitors, ARBs and other agents are included in **Table 2**. In the inpatient cohort, 1731 patients receiving ACE inhibitors and 1560 patients receiving ARBs that were propensity score-matched to patients receiving other anti-hypertensive agents (**eFigure 5**), with covariate standardized differences of <10% after matching (**Figure 1**). The equipoise for comparisons of ACE inhibitors to other drugs, and for ACE inhibitors to ARB were above 0.5, but were lower for the ARB comparisons (**Table 4**).

**Table 2:** Characteristics of Inpatient Study Cohort. The cohort includes patients who were hospitalized with COVID-19.

| Variable | Overall | Antihypertensive Drug Cohorts |  |  | p-value |  |  |
| --- | --- | --- | --- | --- | --- | --- | --- |
|  |  | ACE inhibitor | ARB | Other | ACE inhibitor vs Other | ARB vs Other | ACE inhibitor vs ARB |
| Number of Patients | 7933 | 2361 | 2226 | 3346 |  |  |  |
| **Age, median (IQR)** | 77.0<br>(69.0 - 85.0) | 76.0<br>(68.0 - 83.0) | 76.0<br>(69.0 - 84.0) | 78.0<br>(70.0 - 86.0) | <.001 | <.001 | 0.09 |
| **Age Range** |  |  |  |  |  |  |  |
| 18 to 30 y | 11 (0.1%) | 5 (0.2%) | 3 (0.1%) | 3 (0.1%) | 0.39 | 0.93 | 0.79 |
| 31 to 40 y | 30 (0.4%) | 9 (0.4%) | 6 (0.3%) | 15 (0.4%) | 0.86 | 0.40 | 0.69 |
| 41 to 50 y | 173 (2.2%) | 60 (2.5%) | 47 (2.1%) | 66 (2.0%) | 0.18 | 0.79 | 0.39 |
| 51 to 60 y | 544 (6.9%) | 172 (7.3%) | 177 (8.0%) | 195 (5.8%) | 0.03 | <.001 | 0.43 |
| 61 to 70 y | 1523 (19.2%) | 499 (21.1%) | 415 (18.6%) | 609 (18.2%) | 0.01 | 0.70 | 0.04 |
| 71 to 80 y | 2627 (33.1%) | 822 (34.8%) | 786 (35.3%) | 1019 (30.5%) | <.001 | <.001 | 0.75 |
| > 80 y | 3025 (38.1%) | 794 (33.6%) | 792 (35.6%) | 1439 (43.0%) | <.001 | <.001 | 0.17 |
| Female sex | 4332 (54.6%) | 1171 (49.6%) | 1246 (56.0%) | 1915 (57.2%) | <.001 | 0.37 | <.001 |
| Medicare Advantage | 7296 (92.0%) | 2152 (91.1%) | 1991 (89.4%) | 3153 (94.2%) | <.001 | <.001 | 0.06 |
| **Location** |  |  |  |  |  |  |  |
| Urban | 3574 (45.1%) | 1047 (44.3%) | 1048 (47.1%) | 1479 (44.2%) | 0.94 | 0.04 | 0.07 |
| Rural | 1623 (20.5%) | 488 (20.7%) | 463 (20.8%) | 672 (20.1%) | 0.61 | 0.54 | 0.94 |
| Suburban | 2714 (34.2%) | 822 (34.8%) | 708 (31.8%) | 1184 (35.4%) | 0.68 | 0.01 | 0.03 |
| Unknown | 22 (0.3%) | 4 (0.2%) | 7 (0.3%) | 11 (0.3%) | 0.37 | 0.88 | 0.48 |
| **Race #** |  |  |  |  |  |  |  |
| Caucasian | 4486 (56.5%) | 1352 (57.3%) | 1117 (50.2%) | 2017 (60.3%) | 0.02 | <.001 | <.001 |
| African American | 2181 (27.5%) | 633 (26.8%) | 636 (28.6%) | 912 (27.3%) | 0.73 | 0.30 | 0.19 |
| Hispanic | 209 (2.6%) | 70 (3.0%) | 67 (3.0%) | 72 (2.2%) | 0.06 | 0.05 | 1.00 |
| Asian | 137 (1.7%) | 21 (0.9%) | 73 (3.3%) | 43 (1.3%) | 0.20 | <.001 | <.001 |
| Native American | 8 (0.1%) | 6 (0.3%) | 2 (0.1%) | 0 (0.0%) | 0.01 | 0.31 | 0.33 |
| Other | 156 (2.0%) | 38 (1.6%) | 64 (2.9%) | 54 (1.6%) | 0.93 | <.001 | 0.01 |
| Unknown | 756 (9.5%) | 241 (10.2%) | 267 (12.0%) | 248 (7.4%) | <.001 | <.001 | 0.06 |
| **Geographic Region** |  |  |  |  |  |  |  |
| **Region of Inpatient Facility** |  |  |  |  |  |  |  |
| Northeast | 3335 (42.0%) | 950 (40.2%) | 947 (42.5%) | 1438 (43.0%) | 0.04 | 0.77 | 0.12 |
| South | 2750 (34.7%) | 807 (34.2%) | 829 (37.2%) | 1114 (33.3%) | 0.50 | <.001 | 0.03 |
| Midwest | 1528 (19.3%) | 482 (20.4%) | 378 (17.0%) | 668 (20.0%) | 0.70 | 0.01 | <.001 |
| West | 320 (4.0%) | 122 (5.2%) | 72 (3.2%) | 126 (3.8%) | 0.01 | 0.33 | <.001 |
| **State of Inpatient Facility** |  |  |  |  |  |  |  |
| New York | 1226 (15.5%) | 320 (13.6%) | 386 (17.3%) | 520 (15.5%) | 0.04 | 0.08 | <.001 |
| New Jersey | 796 (10.0%) | 212 (9.0%) | 260 (11.7%) | 324 (9.7%) | 0.39 | 0.02 | <.001 |
| Connecticut | 779 (9.8%) | 238 (10.1%) | 206 (9.3%) | 335 (10.0%) | 0.97 | 0.37 | 0.37 |
| Georgia | 666 (8.4%) | 188 (8.0%) | 208 (9.3%) | 270 (8.1%) | 0.92 | 0.11 | 0.11 |
| Florida | 542 (6.8%) | 141 (6.0%) | 184 (8.3%) | 217 (6.5%) | 0.46 | 0.01 | 0.00 |
| Other | 3924 (49.5%) | 1262 (53.5%) | 982 (44.1%) | 1680 (50.2%) | 0.02 | <.001 | <.001 |
| **Comorbid Conditions** |  |  |  |  |  |  |  |
| Diabetes without complications | 4022 (50.7%) | 1339 (56.7%) | 1237 (55.6%) | 1446 (43.2%) | <.001 | <.001 | 0.45 |
| Myocardial infarction | 425 (5.4%) | 109 (4.6%) | 123 (5.5%) | 193 (5.8%) | 0.06 | 0.75 | 0.18 |
| Chronic heart failure | 2469 (31.1%) | 626 (26.5%) | 656 (29.5%) | 1187 (35.5%) | <.001 | <.001 | 0.03 |
| Chronic pulmonary disease | 2266 (28.6%) | 576 (24.4%) | 588 (26.4%) | 1102 (32.9%) | <.001 | <.001 | 0.12 |
| Peptic ulcer disease | 133 (1.7%) | 36 (1.5%) | 39 (1.8%) | 58 (1.7%) | 0.61 | 0.96 | 0.62 |
| AIDS | 33 (0.4%) | 13 (0.6%) | 6 (0.3%) | 14 (0.4%) | 0.60 | 0.50 | 0.21 |
| Rheumatologic disease | 435 (5.5%) | 91 (3.9%) | 146 (6.6%) | 198 (5.9%) | <.001 | 0.36 | <.001 |
| Diabetes, chronic complications | 3081 (38.8%) | 984 (41.7%) | 963 (43.3%) | 1134 (33.9%) | <.001 | <.001 | 0.29 |
| Metastatic cancer | 146 (1.8%) | 37 (1.6%) | 36 (1.6%) | 73 (2.2%) | 0.12 | 0.16 | 0.99 |
| Hemiplegia or paraplegia | 596 (7.5%) | 189 (8.0%) | 110 (4.9%) | 297 (8.9%) | 0.27 | 0.00 | 0.00 |
| Liver disease, mild | 477 (6.0%) | 120 (5.1%) | 129 (5.8%) | 228 (6.8%) | 0.01 | 0.14 | 0.32 |
| Solid tumor without metastases | 923 (11.6%) | 265 (11.2%) | 252 (11.3%) | 406 (12.1%) | 0.31 | 0.38 | 0.95 |
| Liver disease, moderate to severe | 66 (0.8%) | 17 (0.7%) | 12 (0.5%) | 37 (1.1%) | 0.18 | 0.04 | 0.56 |
| Dementia | 1645 (20.7%) | 481 (20.4%) | 344 (15.5%) | 820 (24.5%) | <.001 | <.001 | <.001 |
| Peripheral vascular disease | 2687 (33.9%) | 755 (32.0%) | 624 (28.0%) | 1308 (39.1%) | <.001 | <.001 | <.001 |
| Renal failure, moderate to severe | 2351 (29.6%) | 592 (25.1%) | 641 (28.8%) | 1118 (33.4%) | <.001 | <.001 | <.001 |
| Cerebrovascular disease | 1744 (22.0%) | 507 (21.5%) | 445 (20.0%) | 792 (23.7%) | 0.06 | <.001 | 0.23 |
| Charlson Score, median (IQR) | 3.0 (2.0 - 5.0) | 3.0 (1.0 - 5.0) | 3.0 (1.0 - 5.0) | 3.0 (2.0 - 6.0) | <.001 | <.001 | 0.77 |
| **Drug Therapy** |  |  |  |  |  |  |  |
| **Antihypertensives** |  |  |  |  |  |  |  |
| Beta blockers | 4277 (53.9%) | 1112 (47.1%) | 1095 (49.2%) | 2070 (61.9%) | <.001 | 0.00 | 0.17 |
| Non-dihydropyridine calcium channel blockers | 3438 (43.3%) | 929 (39.3%) | 959 (43.1%) | 1550 (46.3%) | <.001 | 0.02 | 0.01 |
| Dihydropyridine calcium channel blockers | 2972 (37.5%) | 826 (35.0%) | 848 (38.1%) | 1298 (38.8%) | <.001 | 0.62 | 0.03 |
| Thiazide or thiazide-like diuretics | 1650 (20.8%) | 512 (21.7%) | 702 (31.5%) | 436 (13.0%) | <.001 | 0.00 | 0.00 |
| Loop diuretics | 2400 (30.3%) | 570 (24.1%) | 612 (27.5%) | 1218 (36.4%) | <.001 | 0.00 | 0.01 |
| Centrally acting alpha agonists | 303 (3.8%) | 85 (3.6%) | 98 (4.4%) | 120 (3.6%) | 0.96 | 0.14 | 0.19 |
| Potassium sparing diuretics | 112 (1.4%) | 21 (0.9%) | 22 (1.0%) | 69 (2.1%) | <.001 | 0.00 | 0.85 |
| Mineralocorticoid Aldosterone Antagonists | 435 (5.5%) | 93 (3.9%) | 135 (6.1%) | 207 (6.2%) | 0.00 | 0.90 | 0.00 |
| Renin inhibitors | 0 (0.0%) | 0 (0.0%) | 0 (0.0%) | 0 (0.0%) | nan | nan |  |
| Alpha adrenergic blocking agents | 247 (3.1%) | 70 (3.0%) | 76 (3.4%) | 101 (3.0%) | 0.97 | 0.46 | 0.43 |
| Direct vasodilators | 515 (6.5%) | 110 (4.7%) | 159 (7.1%) | 246 (7.4%) | 0.00 | 0.81 | 0.00 |
| **Place in Therapy** |  |  |  |  |  |  |  |
| First line | 6399 (80.7%) | 2361 (100.0%) | 2226 (100.0%) | 1812 (54.2%) | 0.00 | 0.00 |  |
| Second line | 5478 (69.1%) | 1405 (59.5%) | 1388 (62.4%) | 2685 (80.2%) | 0.00 | 0.00 | 0.05 |
| **Number of antihypertensive classes** |  |  |  |  |  |  |  |
| 1 | 2322 (29.3%) | 442 (18.7%) | 312 (14.0%) | 1568 (46.9%) | 0.00 | 0.00 | 0.00 |
| 2 | 2625 (33.1%) | 850 (36.0%) | 692 (31.1%) | 1083 (32.4%) | 0.00 | 0.33 | 0.00 |
| 3+ | 2986 (37.6%) | 1069 (45.3%) | 1222 (54.9%) | 695 (20.8%) | 0.00 | 0.00 | 0.00 |
| Number, median (IQR) | 2.0 (1.0 - 3.0) | 2.0 (2.0 - 3.0) | 3.0 (2.0 - 3.0) | 2.0 (1.0 - 2.0) | 0.00 | 0.00 | 0.00 |
| **Other Drug Therapies** |  |  |  |  |  |  |  |
| Statins | 4772 (60.2%) | 1528 (64.7%) | 1408 (63.3%) | 1836 (54.9%) | 0.00 | 0.00 | 0.32 |
| Other lipid lowering agents | 423 (5.3%) | 119 (5.0%) | 145 (6.5%) | 159 (4.8%) | 0.66 | 0.01 | 0.04 |
| Oral anticoagulants | 1375 (17.3%) | 384 (16.3%) | 333 (15.0%) | 658 (19.7%) | 0.00 | 0.00 | 0.24 |
| Insulins | 1373 (17.3%) | 461 (19.5%) | 421 (18.9%) | 491 (14.7%) | 0.00 | 0.00 | 0.62 |
| Oral antihyperglycemic agents | 2188 (27.6%) | 820 (34.7%) | 738 (33.2%) | 630 (18.8%) | 0.00 | 0.00 | 0.27 |
| **Follow up** |  |  |  |  |  |  |  |
| Follow up days, median (IQR) | 6.0 (3.0 - 11.0) | 6.0 (3.0 - 11.0) | 6.0 (3.0 - 11.0) | 7.0 (3.0 - 11.0) | 0.81 | 0.92 | 0.75 |
| Days to death, median (IQR) | 6.0 (3.0 - 11.0) | 7.0 (3.0 - 11.0) | 7.0 (3.0 - 13.0) | 5.0 (2.0 - 10.0) | 0.01 | 0.00 | 0.14 |
| Total mortality | 1130 (14.2%) | 319 (13.5%) | 345 (15.5%) | 466 (13.9%) | 0.68 | 0.11 | 0.06 |
| # Race is unknown in all commercially insured members. |  |  |  |  |  |  |  |

### Hospitalizations in Outpatient Cohort

In the outpatient cohort, over a median 30 (IQR 19, 40) days from SARS-CoV-2 testing, patients receiving ACE inhibitors were less frequently hospitalized than those receiving other anti-hypertensive agents (10.7% vs 14.4%, P = 0.03). There was no significant association between ARB therapy and hospitalization rates (12.7% vs 14.4% in individuals with other insurance, P 0.36). In propensity score-matched cohorts (**eFigure 5**), use of neither ACE inhibitors nor ARB was significantly associated with risk of hospitalization (HR: 0.77 [0.53, 1.13], P = 0.18 for ACE inhibitors, and 0.88 [0.61, 1.26], P = 0.48 for ARB, vs other anti-hypertensive agents) (**Figure 1, Table 3**). There were no differences in the a priori chosen falsification endpoints between propensity score-matched populations (**eTable 3**).

**Table 3:** Hazard ratio for Hospitalization among Individuals Testing Positive for SARS-CoV-2 in the Outpatient Setting. Pairwise comparisons from propensity score matched cohorts.

| Comparison Group | Treatment | Control | Matched Treatment | Matched Control | Hazard ratio for hospitalization in outpatient SARS-CoV-2 positive patients (95% CI, P-value) | Equipoise metric |
| --- | --- | --- | --- | --- | --- | --- |
| **Overall population** |  |  |  |  |  |  |
| ACE inhibitor vs Other | 722 | 810 | 441 | 441 | 0.774 (0.530, 1.130); P = 0.18 | 0.68 |
| ARB vs Other | 731 | 810 | 412 | 412 | 0.877 (0.611, 1.258); P = 0.48 | 0.68 |
| ACE inhibitor vs ARB | 722 | 731 | 591 | 591 | 0.913 (0.649, 1.286); P = 0.60 | 0.96 |
| **Medicare Advantage enrollees** |  |  |  |  |  |  |
| ACE vs Other | 581 | 434 | 296 | 296 | 0.614 (0.404, 0.933); P = 0.02 | 0.67 |
| ARB vs Other | 581 | 452 | 283 | 283 | 0.891 (0.586, 1.355); P = 0.59 | 0.68 |
| ACE vs ARB | 452 | 434 | 352 | 352 | 0.879 (0.571, 1.355); P = 0.56 | 0.96 |

There were differences between the association of ACE inhibitors and hospitalization risk across insurance groups (P for interaction, 0.09), with a lower risk of hospitalization in Medicare Advantage patients (HR, 0.61 [0.41, 0.93], P = 0.02) that was not observed in commercially insured individuals (HR, 2.14 [0.82, 5.60], P = 0.12). (**Table 3**).

In propensity score-matched analyses, ARB use was not associated with significantly lower hospitalization risk than individuals receiving other anti-hypertensive agents (HR, 0.88 [0.61, 1.26], P = 0.48) (**Figure 2**). There were no significant differences in hospitalization rates between propensity score matched cohorts of patients receiving ACE inhibitor, compared with ARB (HR 0.91, 0.65 to 1.29; P = 0.60). There were no significant interactions by insurance-type and the association of ARB with outcomes (P-interaction = 0.55)

**Figure 2:**
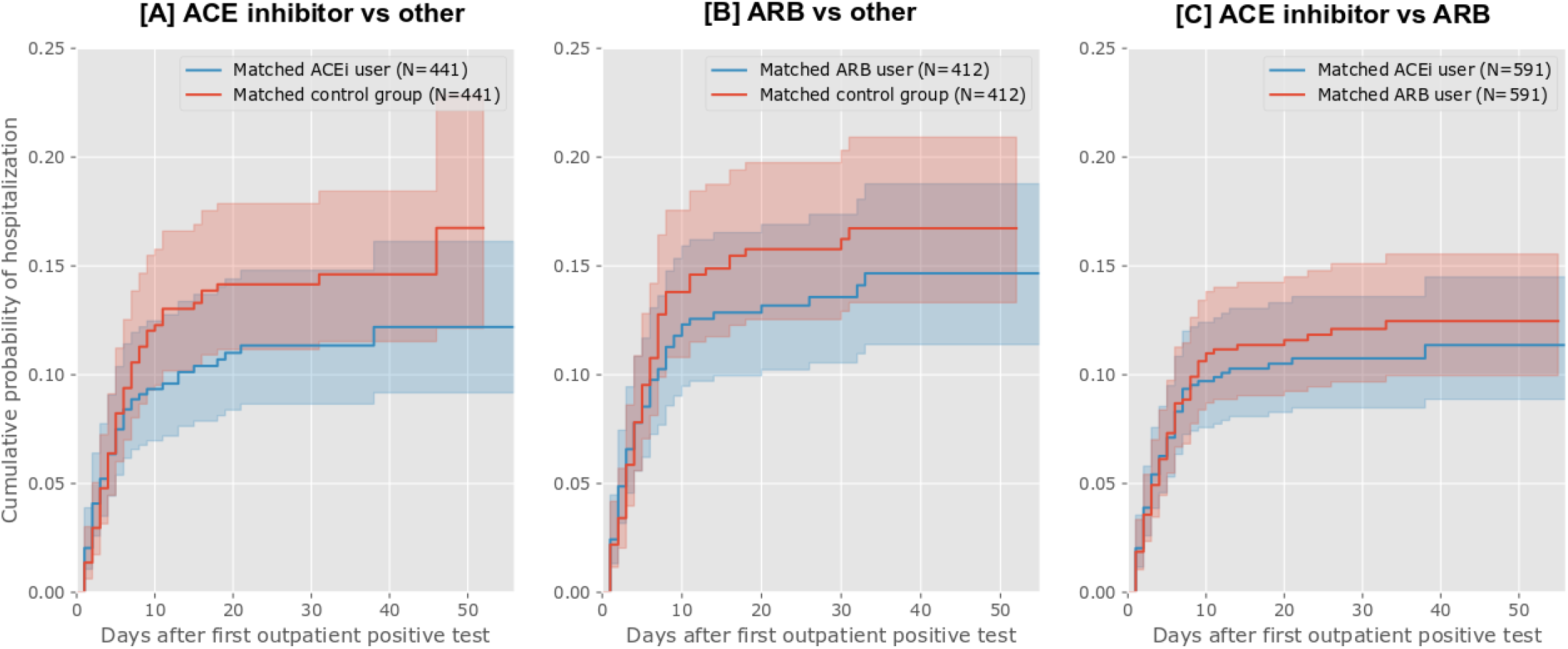
Cumulative event curves for hospitalization among hypertensive patients with a positive SARS-CoV-2 test in the outpatient setting. Plots represents propensity score-matched groups

Among the individuals in the outpatient cohort who were hospitalized, there was no association with ACE inhibitor or ARB use with subsequent in-hospital mortality (**eTable 4**).

### Mortality in the Inpatient Cohort

Of the 7933 patients hospitalized with COVID-19, 1128 (14.2%) died during hospitalization, 4722 (59.5%) were discharged alive, and 2083 (26.3%) were still hospitalized at the end of the observation period. A majority of deaths (90.1%) were among the Medicare Advantage population. The median length of stay (including patients who died as well as discharged alive) for COVID-19 hospitalizations was 6 (IQR: 3 to 11) days, which was similar across patients who died (6 (IQR: 3 to 10) days) or discharged alive (6 (IQR: 3 to 11) days) during the observation period.

Overall, the proportion of COVID-19 patients who died did not differ significantly in those on ACE inhibitor therapy before hospitalization, compared with those on other anti-hypertensive agents (13.5% vs 13.9%, P = 0.68). In the propensity matched-cohort of patients receiving ACE inhibitors before hospitalization, in-hospital mortality was not significantly different than patients on other anti-hypertensive drugs (HR: 0.97 (0.81, 1.16); P = 0.74; **eFigure 5, Table 4**). Similarly, ARB did not have a significantly different risk of mortality compared with other anti-hypertensive agents (1.15 (0.95, 1.38); P = 0.15). There were no significant differences in mortality between patients receiving ACE inhibitors and ARBs in the overall population, without a significant interaction between insurance group and treatment assignment and patient outcome (**Figure 3, Table 4**). These findings were consistent in for our secondary outcome of in-hospital death or discharge to hospice (**eTable 4**). There was also no association between treatment with ACE inhibitor or ARB on hospital length of stay (**eTable 5**).

**Table 4:** Hazard ratio for Mortality in Hospitalized Coronavirus Disease-19 patients. Pairwise comparisons from propensity score matched cohorts.

| Comparison Group | Treatment | Control | Matched | Hazard ratio for mortality in hospitalized for COVID-19 patients<br>(95% CI, P-value) | Hazard ratio for survival to discharge for COVID-19 patients<br>(95% CI, P-value) | Equipoise metric |
| --- | --- | --- | --- | --- | --- | --- |
| **Overall population** |  |  |  |  |  |  |
| ACE inhibitor vs Other | 2360 | 3338 | 1731 | 0.97 (0.81, 1.16); P = 0.74 | 1.03 (0.94, 1.12); P = 0.57 | 0.56 |
| ARB vs Other | 2224 | 3338 | 1560 | 1.15 (0.95, 1.38); P = 0.15 | 1.01 (0.93, 1.11); P = 0.76 | 0.46 |
| ACE inhibitor vs ARB | 2360 | 2224 | 1882 | 0.89 (0.75, 1.05); P = 0.16 | 1.03 (0.95, 1.12); P = 0.47 | 0.95 |
| **Medicare Advantage enrollees** |  |  |  |  |  |  |
| ACE vs Other | 2151 | 3145 | 1580 | 0.89 (0.74, 1.07); P = 0.20 | 1.03 (0.94, 1.13); P = 0.48 | 0.56 |
| ARB vs Other | 1989 | 3145 | 1425 | 1.19 (0.99, 1.44); P = 0.06 | 1.03 (0.93, 1.13); P = 0.58 | 0.46 |
| ACE vs ARB | 2151 | 1989 | 1704 | 0.88 (0.74, 1.04); P = 0.14 | 1.01 (0.92, 1.10); P = 0.89 | 0.95 |

**Figure 3:**
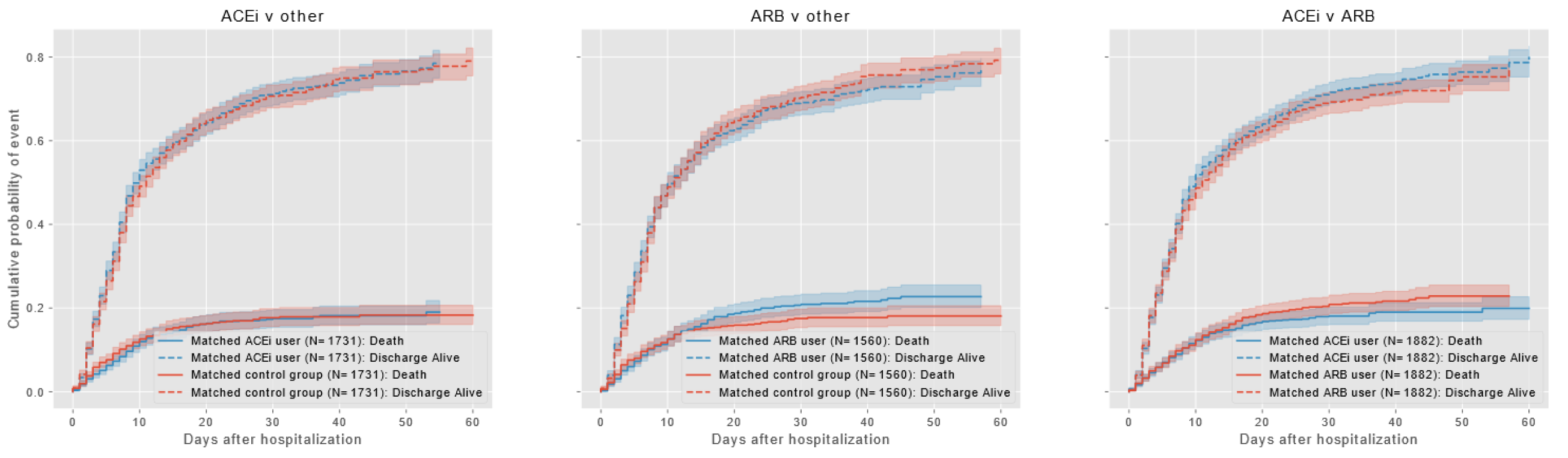
Cumulative event curves for in-hospital mortality among hypertensive patients with hospitalization for COVID-19. Plots represents propensity score-matched groups.

Sensitivity analyses that focused on individuals receiving at least one first line antihypertensive agent in the control group, and that varied the covariate adjustment strategies were consistent with the primary analysis (**eTables 6-8**). The 100 iterations of propensity score matching algorithm found a lower risk of hospitalization with ACE inhibitors across all iterations, with a P-value of less than 0.05 in 97 of 100 iterations in the Medicare Advantage population.

## DISCUSSION

In this first national study of ACE inhibitors and ARBs in outpatients testing positive for SARS-CoV-2, we found that overall these drugs did not confer additional risk or benefit. However, among those in Medicare Advantage, ACE inhibitors were associated with an almost 40% significantly lower risk of hospitalization for COVID-19, an effect not observed with ARBs. Among those hospitalized for COVID-19, we did not find a benefit or a harm of these medications. Collectively, the findings do not support a change to the current use of these medications but, given the lack of effective therapies to mitigate the harm of the virus, this study does provide a rationale for testing the use of ACE inhibitors in older patients to reduce the risk of severe SARS-CoV-2 infection.

Our design has several attributes that should provide confidence about the findings even though it is observational. Our study was restricted to people with hypertension who were receiving at least one anti-hypertensive agent, thereby limiting our assessment to individuals receiving treatment for the same chronic illness, and therefore, equally likely to seek care for healthcare needs for COVID-19. In all analyses, we explicitly compared individuals with equipoise for receiving either drug treatment. Moreover, we did not find any evidence of confounding by disease severity in choice of therapy in our assessment of a priori defined falsification endpoints. Further, our study included individuals from across the United States, thereby limiting the effect of hospital or regional care practices that may bias an evaluation of treatment effects.

Our observations extends the prior evidence of supporting safety of ACE inhibitor treatment in COVID-19.^4,7,9,22^ We show the safety of these agents in outpatient SARS-CoV-2 infected individuals, which complements the studies of those who were hospitalized.^4,7,22^ Also, ours is a national study, which complements the single center or single region studies.^23^

Our study does have an intriguing finding. In the subgroup of individuals enrolled in Medicare advantage, we find that ACE inhibitors were associated with a significantly lower risk of hospitalization following an infection with SARS-CoV-2 in the outpatient setting. These patients are older, more frequently have comorbidities, and were more vulnerable to severe COVID-19 disease.^4,7^ These findings are particularly relevant as the Medicare Advantage group represented over 90% of all COVID-19 hospitalizations in our inpatient cohort. Also, interestingly, we did not find the same association for ARBs despite clinical equipoise in the use of these drugs. Our findings are consistent with prior evidence from randomized clinical trials that a reduced risk of pneumonia with ACE inhibitors that is not observed with ARB.^5^

Our study of in-hospital outcomes adds to the literature on studies that have reached contrasting conclusions regarding the role of ACE inhibitor therapy and in-hospital mortality among hospitalized COVID-19 patients. We did not find a significant association with mortality, consistent with others that have not found such an association.^4,23,24^ However, our findings contrast with certain studies that have found lower mortality in hospitalized COVID-19 patients treated with ACE inhibitors.^7,22^ Notably, the studies that have evaluated mortality risk with COVID-19 before have not consistently been designed to detect potential causal association of drug therapy with outcomes,^7^ relied on case control designs,^22^ pursued potentially biased assessment by using comparators not receiving any therapy,^4,7^ or are based on data from single health centers.^4,23^ Moreover, some have studied both ACE inhibitors and ARBs together despite potentially different mechanisms and effects on patient outcomes.^5^

There some rationale for why there may be a specific ACE inhibitor effect. There is preclinical evidence that suggests a possible protective role for ACE inhibitors in COVID-19. ACE inhibitors, but not ARBs, are associated with the upregulation of ACE-2 receptors.^3,25^ These receptors modulate the local renin-angiotensin-aldosterone system interactions in the lung tissue.^26^ The presence of ACE2 receptors, therefore, exerts a protective effect against the development of acute lung injury in infections with SARS coronaviruses, which lead to dysregulation of these mechanisms and endothelial damage.^27,28^ Further, our observations do not support a theoretical concerns of adverse outcomes due to enhanced virulence of SARS coronaviruses due to overexpression of ACE2 receptors in cell cultures – an indirect binding site for these virues.^29^ However, the pathophysiological effects of ACE inhibitors in SARS-CoV2 infection and the development of clinical disease are likely complex and require dedicated investigation.

Our study has important implications for 4 ongoing randomized trials as none of them align with the observations of our study.^6^ Of the 4 trials, 3 are testing the use of ACE inhibitors or ARBs in the treatment of hospitalized COVID-19 patients, and 1 is using a 10-day course of ARBs after a positive SARS-CoV-2 test to prevent hospitalization.^6^ However, our study suggests that the most appropriate strategy to test in a trial would be the prophylactic use of ACE inhibitor to prevent hospitalization, which is not being tested in any current study.

The findings of our study should be interpreted in light of the following limitations. First, the study is observational, and despite robust methods, and explicit assessments of residual confounding, understanding the potential protective role of ACE inhibitors in COVID-19 requires a dedicated randomized controlled trial. Second, we do not know the proportion of patients receiving these antihypertensive agents that continued to be treated with these drugs during the illness and the association of their continued use or cessation with patient outcomes. Third, all included data elements are contingent upon individuals seeking care for that ailment or filling a medication using their insurance provider and would not be captured if they chose to self-pay. However, we do not expect that any sizeable proportion of insured individuals would defer insurance coverage for their care. Fourth, we cannot account for differences in timing of presentation of patients relative to their symptom onset. However, we limited the effect of differential presentation by patients across exposure groups by focusing on patients receiving treatment for the same medical comorbidity, i.e. hypertension, and only varying the class of drugs. Moreover, we included patients across the US and accounted for clustering of patients, thereby limiting the effect of local practice patterns that may affect hospitalization thresholds. Therefore, it is unlikely patient’s care seeking behavior would be affected by knowledge of their underlying disease. Finally, while our analyses of hospitalization use all available evidence for disease severity, we do not have granular details on real-time inpatient treatment of patients with COVID-19 and whether certain presentation or care characteristics are associated with in-hospital outcomes. Instead, our study evaluates the association of only pre-hospital factors with patient outcomes during hospitalization.

In conclusion, the use of ACE inhibitors and ARBs was not associated with the risk of hospitalization or mortality among those infected with SARS-CoV-2. However, there was a nearly 40% lower risk of hospitalization with the use of ACE inhibitors in the Medicare population. This finding merits a clinical trial to evaluate the potential role of ACE inhibitors in reducing the risk of hospitalization among older individuals, who are at an elevated risk of adverse outcomes with the infection.

## Data Availability

The data are proprietary and are not available for open sharing due to restrictions on our data use agreement

